# Maternal Colonization, Perinatal Exposure, and Neonatal Acquisition of Resistant Enterobacterales

**DOI:** 10.1101/2024.06.28.24309630

**Authors:** Leena B. Mithal, Alima Sajwani, Abigail Aron, Aspen Kremer, Jack T. Sumner, Valeria C. Castro Manzano, Grayson Donnelly, Andrew D. Watson, Weitao Shuai, Emily S. Miller, Joel B. Fisher, Erica M. Hartmann, Mehreen Arshad

## Abstract

**Background:** Multi-drug resistant Enterobacterales, in particular with ceftriaxone resistance (CefR-E), are globally prevalent. Pregnant people colonized with resistant Enterobacterales are at risk of perinatally transmitting to neonates; infections with CefR-E strains are associated with higher mortality, morbidity and health care costs. This study aimed to estimate the rate of gut colonization of AmpR-E (ampicillin resistant Enterobacterales) and CefR-E in a population of healthy mother-infant dyads in the Chicago area and investigate the genetic characteristics of CefR-E.

**Methods:** Pregnant persons anticipating vaginal birth at two Chicago area hospitals were enrolled. Patients with preterm delivery (<35 weeks), fever during labor, cesarean delivery, exposure to antibiotics in 3rd trimester, and neonatal intensive care were excluded. Pregnancy and birth history were obtained from parent and medical record. Maternal vaginal and rectal swabs, and infant stool samples were collected and screened for resistance by plating on MacConkey with ceftriaxone. Whole genome sequencing (WGS) and analysis was conducted on the CefR-E isolates.

**Results:** Between July 2020 and April 2022, 261 mother-infant dyads were enrolled with 184 maternal samples and 157 infant stool samples analyzed. Rate of maternal AmpR-E gut colonization was 97% (179/184). Rate of maternal and infant CefR-E gut colonization was 14% (26/184) and 7% (11/157) respectively. Perinatal transmission of CefR-E was 50% (11 of 22) positive mothers with available infant sample). No clinical variables were significantly associated with maternal CefR-E colonization or perinatal transmission. Type of infant nutrition (breastmilk) was significantly associated with decreased perinatal transmission of AmpR-E to infant (p=0.015). WGS of the CefR-E showed that 19/42 isolates were *E. coli*, and transmission primarily occurred from mothers colonized with *E. coli*.

**Conclusions:** This study demonstrates that a sizable minority of pregnant persons are colonized with CefR-E in the U.S., a higher burden than previously reported for developed countries. Enterobacterales appear adept in perinatal transmission, with half of infants born to colonized pregnant persons harboring CefR-E in first week of life.

## Introduction

Gram-negative bacteria such as *Escherichia coli* are a leading cause of neonatal infections^1-3^. Third generation cephalosporins make up the backbone of neonatal sepsis guidelines across the world^4^. Over the last two decades there has been an annual increase of 3.2% in the incidence of pediatric bloodstream infections with Ceftriaxone Resistant Enterobacterales (CefR-E), the majority of which are producers of Extended Spectrum Beta Lactamases (ESBLs)^5^. In developing countries, mortality is higher in neonates and infants infected with CefR-E likely attributed to delays in appropriate therapy^6^. However, even in developed countries, CefR-E infections significantly increase neonatal morbidity, length of hospital stay and health-care costs^7^. The disability-adjusted life-years (DALYs) for infants <1 year infected with CefR-E is significantly higher than any other age group^8^.

In women of reproductive age, Enterobacterales, especially extra-intestinal pathogenic *E. coli* (ExPEC) that colonize the gut, are part of the microbial reservoirs in perianal, periurethral and vaginal areas, and are thus proximal to the emerging newborn during vaginal birth^9^. Maternal colonization rates with CefR-E vary widely from 2.9% in Norway to 17.6% in Africa^10,11^, but can be passed on to infants by perinatal transmission, colonize the infant gut, and subsequently increase the risk of invasive infections in infants in the neonatal intensive care unit or contribute to the community reservoir of these strains in otherwise healthy infants^11,12^.

Given the increase in pediatric infections with antimicrobial resistant bacteria within the U.S. as well as the rise in community-acquired infections with resistant pathogens, we aimed to determine the molecular epidemiology of maternal and neonatal colonization with CefR-E in otherwise healthy pregnant people and newborn infants at two hospitals in Chicago, IL, report rate of perinatal transmission, and present the genomic characteristics of the isolated CefR-E.

## Methods

### Patient Enrollment

As part of an observational prospective cohort study, pregnant persons were approached during their labor and delivery hospitalization and enrolled prior to delivery between January 2020 to April 2022. We identified pregnant persons anticipating a vaginal delivery at Northwestern Medicine Prentice Women’s Hospital (PWH) and Northwest Community Hospital (NCH) in Chicago and Arlington Heights, IL, U.S. respectively. Pregnant persons who presented with preterm delivery (<35 weeks), fever during labor, planned cesarean delivery, and recorded antibiotic use in the third trimester were excluded from the study and not approached. If a subject was enrolled and pregnancy subsequently resulted in unplanned cesarean delivery, infant neonatal intensive care unit admission, or infant postnatal antibiotic receipt before 24 hours of life, their data were excluded from this study (**Figure 1A**). A survey was administered at the time of enrollment to obtain travel and medical history, including chronic medical conditions, daily medications, country of origin, and travel outside of the U.S. in the two years prior to enrollment. Additional clinical data were abstracted from medical records, including information regarding the infant’s delivery, hospital course, and infant nutrition. Maternal vaginal and rectal swabs were collected by the obstetric care providers prior to delivery. Infant stool samples were collected from a soiled diaper between 24-hours of life and discharge from the hospital. A second infant stool diaper was collected between 7-10 days of life. This study was approved by the Institutional Review Board of Northwestern University (STU#2020-3331) and all study activities were in accordance with the Declaration of Helsinki.

**Figure 1.**
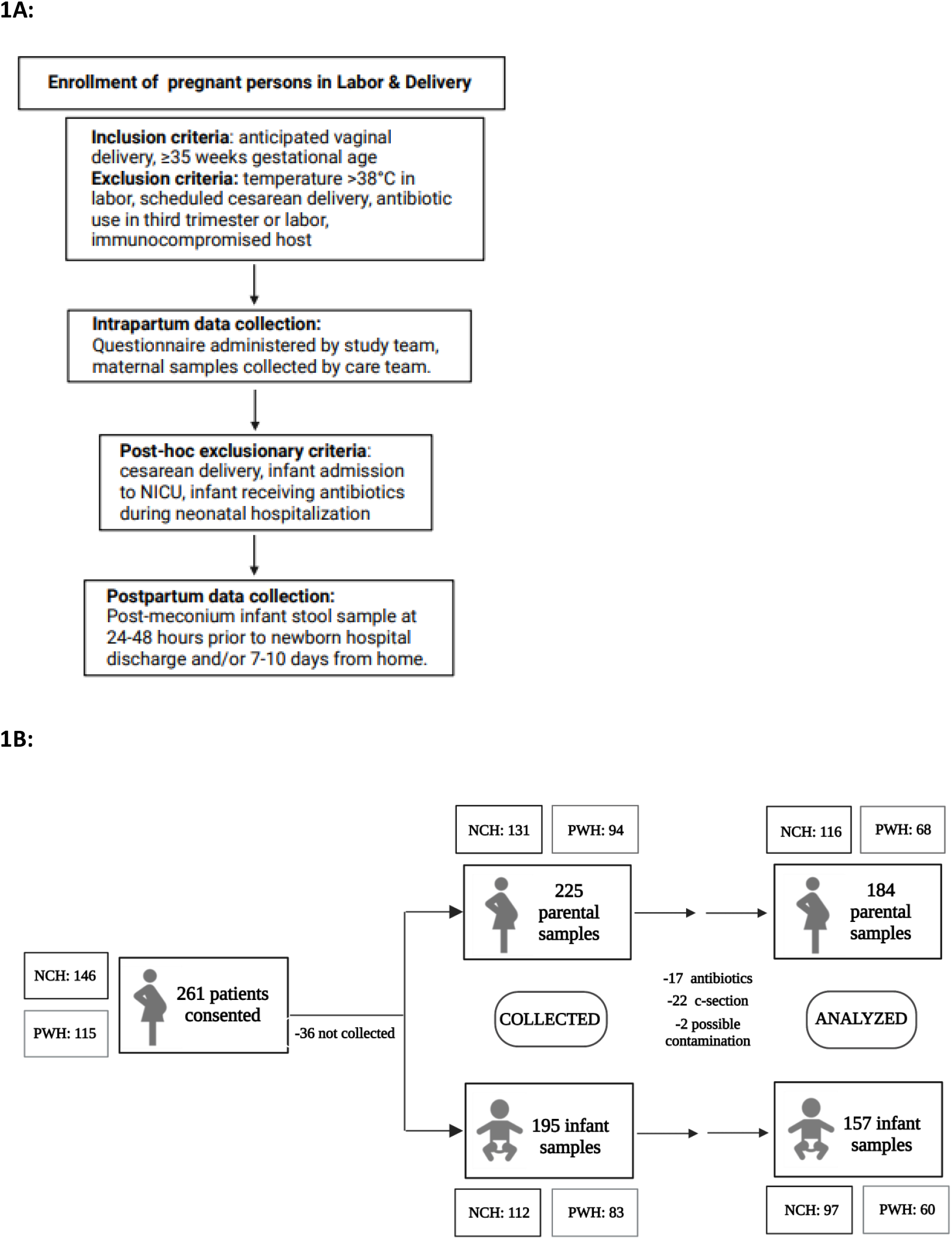
Patient cohort: enrollment and maternal/infant samples. Northwest Community Hospital (NCH), Northwestern Medicine Prentice Women’s Hospital (PWH). Final “n” for analysis was 184 maternal and 157 infant samples. Infant samples were collected during hospital admission (1-2 days of life) and/or at 7-10 days of life.

### Sample processing

Maternal vaginal and rectal swabs that were collected during labor were immersed in culture tubes containing LB, ampicillin, and vancomycin. A pea-sized amount of stool was collected from soiled infant diapers and immersed in the culture tubes mentioned above. All LB samples were incubated overnight for 14-16 hours at 37°C in a shaking incubator and screened for AmpR-E and CefR-E using MacConkey agar and the relevant antibiotic marker (ampicillin or ceftriaxone). Plates were incubated overnight and growth of colonies on the plates assessed.

### Genomic DNA extraction and sequencing

For both maternal and infant stool samples that grew in the presence of ceftriaxone, single bacterial colony isolates were sequenced to determine 1) if and which CefR-E strains were isolated and 2) if the same CefR-E strain was transmitted from mother to infant. Genomic DNA was extracted using the Wizard® Genomic DNA Purification Kit (Promega). The isolated DNA was subjected to sequencing library preparation. For this step, 1 ng DNA was used to start the process using the Illumina Nextera XT DNA Sample Preparation Kit. DNA tagmentation, PCR amplification of tagmented DNA, PCR reaction cleanup, and library quantification and normalization were done. The Illumina MiSeq sequencer was used to sequence the libraries to produce paired-end 300bp reads.

### Genomic analysis

Raw paired reads were trimmed and quality controlled using FastP^13^ before assembling “clean” reads with SPAdes^14^. Putative gene models were identified using prodigal^15^ which were then functionally annotated with eggNOG-mapper^16^. The taxonomic lineage of each assembled genomes was assigned using the Genome Taxonomy Database Tool Kit (GTDB-tk)^17^, and a phylogenetic tree was reconstructed using PhyloPhlAn2^18^ from putative protein coding genes. Finally, this tree, with GTDB-tk annotations (i.e., species names), was visualized using GraPhlAn^19^.

To identify putative transmission events between a mother and their infant, we quantified single nucleotide variants (SNVs) among all pairs of study isolates collected from mothers and their infants using bioinformatics analyses and clonality definitions as previously described^37^. Additionally, traditional 7-gene multilocus sequence typing (MLST) as well as core genome MLST was performed using Enterobase to assess strain relatedness and assign sequence type^38^.

### Identification of resistance, virulence, and metabolic genes

Pathosystems Resource Integration Center (PATRIC)^20^ was used to perform a BLAST-based comparison using PATRIC’s collection of antibiotic resistance proteins and the externally curated Comprehensive Antibiotic Resistance Database (CARD)^21^. Similarly, the Comprehensive Genome Analysis tool within PATRIC was used to identify virulence factors that are known to play a role in adhesion to gut epithelium and therefore play a role in gut colonization (curli fibers, fimbriae, pilus and dispersin aggregated proteins)^22^. This tool also uses BLAST-based comparisons of annotated genome with several databases including PATRIC virulence factor database (PATRIC_VF), and the externally curated databases Victors (a web-based resource for virulence factors in human and animal pathogens, and Virulence Factor DataBase (VFDB). For all BLAST based comparisons the default settings within PATRIC were used, with maximum hits of 50 and an E-value threshold of 10.

### Statistical analysis

Demographics and clinical characteristics including age, race, ethnicity, gestational or preexisting diabetes, medications, type of infant nutrition, and country of birth were reported and compared between groups with and without AmpR Enterobacterales colonization and with and without CefR Enterobacterales colonization with Chi-square and Fisher’s Exact Tests as appropriate for categorical variables. Rates of perinatal transmission were calculated. For the purpose of reporting infant acquisition of resistant strains we considered a sample to be positive if either of the samples (24-48 hour or 7-10 day) grew Ampicillin or Ceftriaxone resistant bacteria, respectively. Given samples were collected within the first week of life, if infant was CefR-E positive and maternal swab was not positive, rates with calculated with the more biologically plausible assumption of maternal CefR-E colonization. Sensitivity analysis calculation was also conducted for perinatal transmission. For all analyses, statistical significance was defined as p <0.05, without correction for multiple comparisons. Analyses were conducted using STATA/IC version 16.0 (Statacorp).

## Results

### Patient enrollment

We consented 261 pregnant patients in total (**Figure 1B**). Of these 146 were from NCH and the remaining 115 were from the PWH. Rectal and vaginal samples were collected from a total of 225 maternal samples between the two sites. Stool samples from the first 24-48 hours of life and/or the at 7-10 days of life were collected from 195 infants. Sample analysis was ultimately done on samples from 184 mothers and 157 infants, due to exclusion of mothers who received antibiotics during labor after enrollment (n=17), Caesarian delivery (n=22), or possible contamination of samples (n=2).

### Demographic data

The mean maternal age at time of delivery was 32.9 years (SD 4.5). The cohort was 78% white, 13% Asian, 6% Other/Unknown, and had 1 Black and 1 American Indian patient. Seventeen patients (9%) were Latinx ethnicity. Eight percent had gestational diabetes and 2% had preexisting diabetes before pregnancy. Probiotic use was reported in 11% and acid-blocking medicines such as H2 inhibitors or proton pump inhibitors were used by 19% of the cohort. Twenty-three percent was born outside of the U.S. The majority of infants were receiving exclusive breastmilk at the time of hospital discharge (64%), 7% received formula only, and 30% had intake of both breastmilk and formula (**Table 1**).

**Table 1:** Characteristics of Mother-Infant Dyads by Colonization and Perinatal Transmission.

| n (%) | Total cohort<br>(n=184) | Maternal<br>colonization<br>with<br>AmpR-E<br>(n=179) | Maternal<br>colonization<br>with CefR-E<br>(n=26) | No maternal<br>CefR-E<br>colonization<br>(n=158) | Infant<br>colonization<br>with AmpR-E<br>(n=117/153) | Infant<br>colonization<br>with CefR-E<br>(n=11/22) | *p-value*<br>Maternal<br>CefR-E<br>colonization | *p-value*<br>Amp-R E<br>transmission | *p-value*<br>CefR-E<br>transmission |
| --- | --- | --- | --- | --- | --- | --- | --- | --- | --- |
| **Age**<br>mean (std) | 32.9 (4.5) | 33.1 (5.0) | 33.9 (4.5) | 32.9 (5.0) | 33.3 (4.3) | 33.5 (3.0) |  |  |  |
| **Race** |  |  |  |  |  |  | 0.168 | 0.382 | 0.453 |
| Black | 1 (0.5%) | 1 (0.6%) | 0 | 1 (0.6%) | 1 (0.9%) | 0 |  |  |  |
| White | 148 (78%) | 143 (80%) | 17 (65%) | 131 (83%) | 91 (78%) | 8 (73%) |  |  |  |
| Asian | 23 (13%) | 23 (13%) | 6 (23%) | 17 (11%) | 19 (16%) | 1 (9%) |  |  |  |
| American Indian<br>or Alaskan<br>Native | 1 (0.5%) | 0 | 0 | 0 | 0 | 0 |  |  |  |
| Other | 11 (6%) | 11 (6%) | 3 (12%) | 8 (5%) | 6 (5%) | 2 (18%) |  |  |  |
| **Latina/Hispanic** | 17 (9%) | 16 (9%) | 3 (12%) | 14 (9%) | 10 (9%) | 2 (18%) | 0.565 | 0.732 | 1.0 |
| **Preexisting or**<br>**gestational**<br>**diabetes** | 19 (10%) | 18 (10%) | 5 (19%) | 14 (9%) | 13 (11%) | 3 (27%) | 0.107 | 0.812 | 0.586 |
| **Probiotics** | 20 (11%) | 19 (11%) | 3 (12%) | 17 (11%) | 9 (8%) | 1 (9%) | 1.0 | 0.175 | 1.0 |
| **Acid-blocking**<br>**medication** | 35 (19%) | 34 (19%) | 3 (12%) | 32 (20%) | 21 (18%) | 1 (9%) | 0.294 | 0.333 | 1.0 |
| **Nutrition** |  |  |  |  |  |  |  | **0.015** | 0.12 |
| Breastmilk | 114 (64%) | 110 (63%) | 17 (68%) | 97 (63%) | 63 (55%) | 8 (73%) |  |  |  |
| Formula | 12 (7%) | 11 (6%) | 2 (8%) | 10 (6%) | 10 (9%) | 2 (18%) |  |  |  |
| Both | 53 (30%) | 53 (30%) | 6 (24%) | 47 (31%) | 41 (36%) | 1 (9%) |  |  |  |
| **Born outside**<br>**U.S.** | 43 (23%) | 43 (24%) | 9 (35%) | 34 (22%) | 30 (26%) | 3 (27%) | 0.321 | 0.295 | 0.375 |

Comparisons were by Chi-square and Fisher's Exact Tests, as appropriate. For purpose of data analysis, if infant stool was positive for CefR-E, mother was considered CefR-E positive even if rectal swab did not yield CefR-E in culture.

### Maternal and Infant Gut Colonization Rates and Perinatal Transmission

In our cohort, 97% of mothers were colonized with AmpR-E, and a further 76% of the infants were carriers of AmpR-E in the first week of life. Among the same cohort, 11% of the maternal rectal samples and 7.2% of the infant stool samples were positive for CefR-E. The rate of maternal CefR-E colonization at our suburban site was 16/116 (14%) and at urban hospital 10/68 (15%).

Twenty mothers were positive for CefR-E on vaginal/rectal culture (20/184, 11% plus an additional 6 dyads for which infant stool was culture positive for CefR-E and maternal CefR-E colonization was presumed). Fourteen percent (26/184) of mothers were CefR-E colonized and perinatal transmission rate was 50% (11/22) (**Figure 2**). In sensitivity analysis, considering the 20 maternal samples that were positive in culture, 16 had infant samples available and 5 were positive yielding a most conservative perinatal transmission rate of 31%.

**Figure 2.**
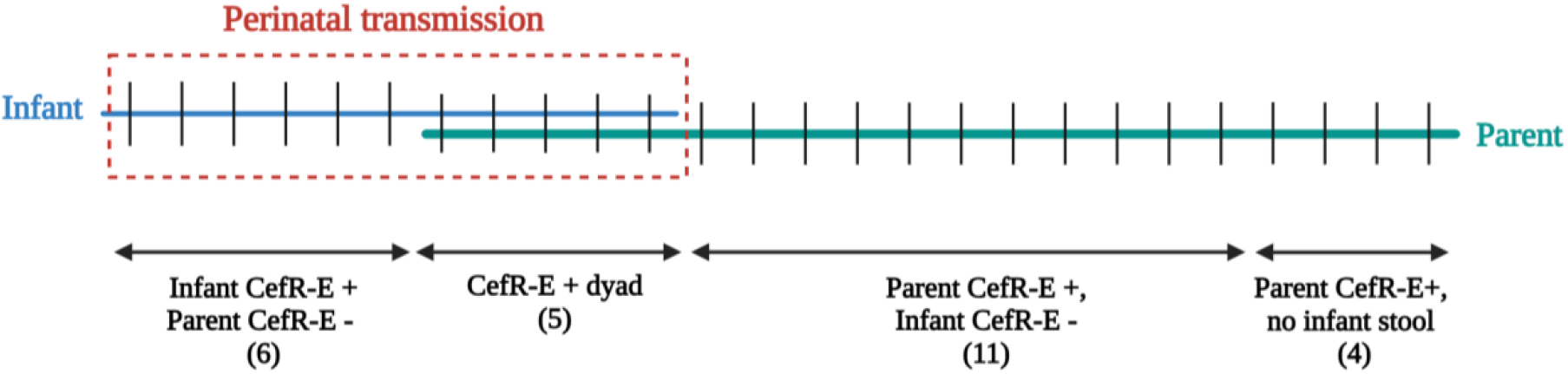
Mother and infant colonization with CefR-E. Red dotted line represents perinatal transmission of CefR-E. If infant stool was positive for CefR-E, perinatal transmission was assumed from CefR-E positive mother despite maternal rectal swab negative for CefR-E in culture (n=6).

### Clinical characteristics associated with colonization and perinatal transmission

Race, ethnicity, diabetes mellitus, probiotic use, acid-blocking medications, and birth outside of the U.S. were not significantly associated with CefR-E colonization in mothers. Similarly, these factors were not significantly associated with infant colonization of AmpR-E or CefR-E (**Table 1**). Type of infant nutrition was significantly associated with perinatal transmission of AmpR-E to infant (p=0.015). Among infants who acquired AmpR-E, 55% were exclusively breastfed, 9% formula fed, and 36% both breastmilk and formula compared to infants who did not acquire AmpR-E who were 80% exclusively breastfed (28/35), none exclusively formula fed, and 20% received both. No clinical variables were significantly associated with maternal CefR-E colonization or perinatal transmission, although our power to detect differences was limited given absolute CefR-E numbers.

### Phylogenetic analysis of isolated CefR-E

Figure 3. shows the phylogenetic tree of CefR-E isolated from mothers and infants, 14 were found to be *Escherichia spp*, 11 were *Enterobacter spp*, 7 were *Citrobacter spp*, 3 were *Klebsiella spp*, and 1 was of *Proteus spp*. The mother-infant dyad NCH0126 were noted to have two different CefR-E species: *Klebsiella pneumoniae* in the mother and *Enterobacter cloacae* in the infant. Two different CefR-E species were also found in the PWH0104 maternal sample. Interestingly both the *E. coli* and *Proteus mirabalis* species carried the same beta lactamases. Two different morphologies (pink vs. white) of *E. coli* were noticed in the NC0104 dyad, but both were found to be ST131 with a similar resistance gene profile.

The distribution of the genus, species and sequence type where they could be determined, along with the beta-lactamase gene is shown in **Table 2**. Notable sequence types within *E. coli* were ST131, ST10 and ST69 all of which have been associated with ESBL production.

**Table 2.** Resistant genes across mother-infant dyads.

| Strain Type | Genus | Species | Strain | B-lactamase genes |
| --- | --- | --- | --- | --- |
| NCH0002 | *Enterobacter* | *cloacae* |  | ACT/MIR family |
| NCH0009 | *Enterobacter* | *cloacae* |  | CHM family, MIR-10 |
| NCH0012 | *Enterobacter* | *cloacae* |  | CHM family, MIR-10 |
| NCH0016 | *Escherichia* | *coli* | ST10 | CTX-M-15, BLaEC family |
| NCH0018 | *Enterobacter* | *cloacae* |  | ACT/MIR family, ampC |
| NCH0023 | *Escherichia* | *coli* | ST131 | CTX-M-15, BLaEC family |
| NCH0024 | *Enterobacter* | *cloacae* |  | ACT/MIR family |
| NCH0031 | *Escherichia* | *coli* | ST10 | BLaEC family |
| NCH0033 | *Enterobacter* | *cloacae* |  | ACT/MIR family |
| NCH0047 | *Escherichia* | *coli* | ST12991 | CTX-M-15, BLaEC family, TEM-12 |
| NCH0049 | *Escherichia* | *coli* | ST69 | CTX-M-15, BLaEC family, TEM-12 |
| NCH0076 | *Escherichia* | *coli* | ST10 | CTX-M-15, BLaEC family, LAP family, TEM-12 |
| NCH0104 - mother | *Escherichia* | *coli* | ST131 | CTX-M-15, BLaEC family, OXA-1 |
| NCH0104 - infant | *Escherichia* | *coli* | ST131 | CTX-M-15, BLaEC family, OXA-1 |
| NCH0112 - mother | *Escherichia* | *coli* | ST349 | BLaEC family, DHA/MOR family |
| NCH0112 - infant | *Escherichia* | *coli* | ST349 | BLaEC family, DHA/MOR family |
| NCH0126 - mother | *Klebsiella* | *pneumonia* |  | OKP-B family |
| NCH0126 - infant | *Enterobacter* | *cloacae* |  | ACT/MIR family |
| NCH0136 | *Citrobacter* | *freundii* |  | CMY-4 |
| NCH0137 | *Citrobacter* | *sp.* |  | CMY-4 |
| NCH0138 | *Escherichia* | *coli* | ST410 | CTX-M-15, BLaEC family, LAP family, TEM-12, CMY-4, OXA-1 |
| NCH0141 | *Escherichia* | *coli* | ST162 (ST469 complex) | CTX-M-15, BLaEC family |
| PWH0017 | *Enterobacter* | *cloacae* |  | ACT family |
| PWH0025 | *Escherichia* | *coli* | ST131 | CTX-M-15, OXA-1 |
| PWH0028 | *Escherichia* | *coli* | ST6316 (ST446 complex) | BLaEC family |
| PWH0031 - mother | *Escherichia* | *coli* | ST569 | CTX-M-15, BLaEC family |
| PWH0031 - infant | *Escherichia* | *coli* | ST569 | CTX-M-15, BLaEC family |
| PWH0041 | *Enterobacter* | *cloacae* |  | ACT/MIR family |
| PWH0050 | *Klebsiella* | *oxytoca* |  | OXY family |
| PWH0060 | *Citrobacter* | *freundii* |  | CMY-4 |
| PWH0062 | *Citrobacter* | *sp.* |  | CMY-4 |
| PWH0073 | *Klebsiella* | *aerogenes* |  | OXYR |
| PWH0074 | *Citrobacter* | *sp.* |  | CMY-4 |
| PWH0086 | *Citrobacter* | *freundii* |  | CMY-4 |
| PWH0094 | *Enterobacter* | *cloacae* |  | ACT/MIR family |
| PWH0104 -<br>mother | *Proteus* | *mirabilis* |  | CTX-M-15, BlaEC family, OXA-1 |
| PWH0104 -<br>mother | *Escherichia* | *coli* | ST69 | CTX-M-15, BlaEC family, OXA-1 |

**Figure 3.**
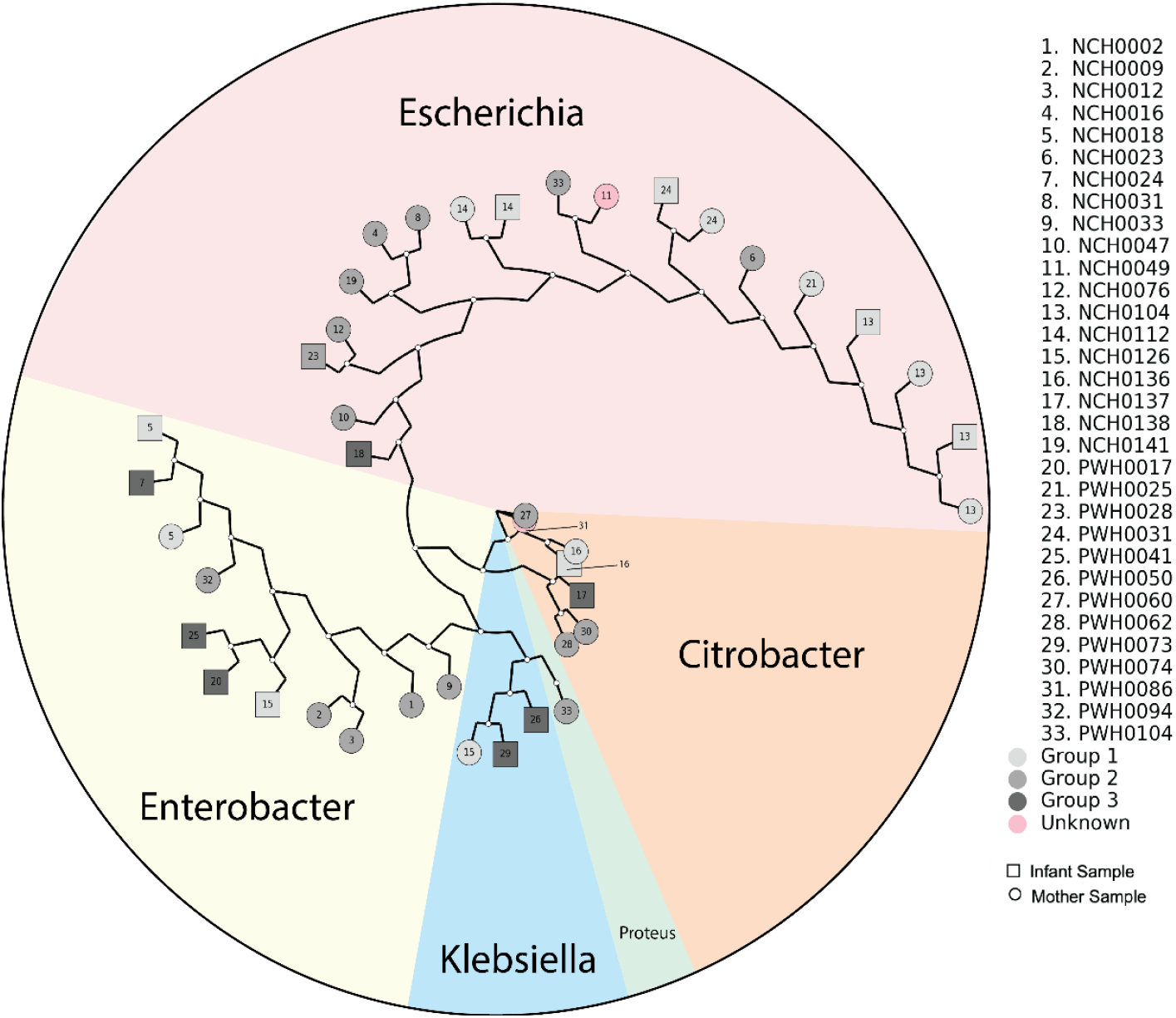
Whole genome sequencing of the ESBL Enterobacterales isolates. WGS showed that 45% were *E. coli* (19/42), followed by *Enterobacter* (11), *Citrobacter* (7), and *Klebsiella* (3). Group number is indicated by grayscale color as indicated in legend. Group 1 – CefR-E+ dyads; Group 2 – CefR-E+ mother, CefR-E-infant; Group 3 – CefR-E+ infant, CefR-E-mother; Unknown – CefR-E+ mother, sample not collected for infant.

### Resistance gene profile of strains

As shown in **Table 2**, almost all *E. coli* species carried ESBL genes with the CTX-M genes being the most common, followed by the OXA and TEM ESBLs. Genes conferring resistance against ceftriaxone amongst *Enterobacter* spp included ACT/MIR and CHM families. All 6 *Citrobacter* spp were associated with the cephalosporinase CMY-4, whereas *Klebsiella* carried the OXY-family beta-lactamase. **Figure 4** shows the frequency of non-beta lactamase genes found in the CefR-E isolates. All CefR-E isolates were found to carry resistance genes against multiple classes of antibiotics. **Supplemental Table** shows the resistance genes associated with the different non-beta lactam classes of antibiotics.

**Figure 4.**
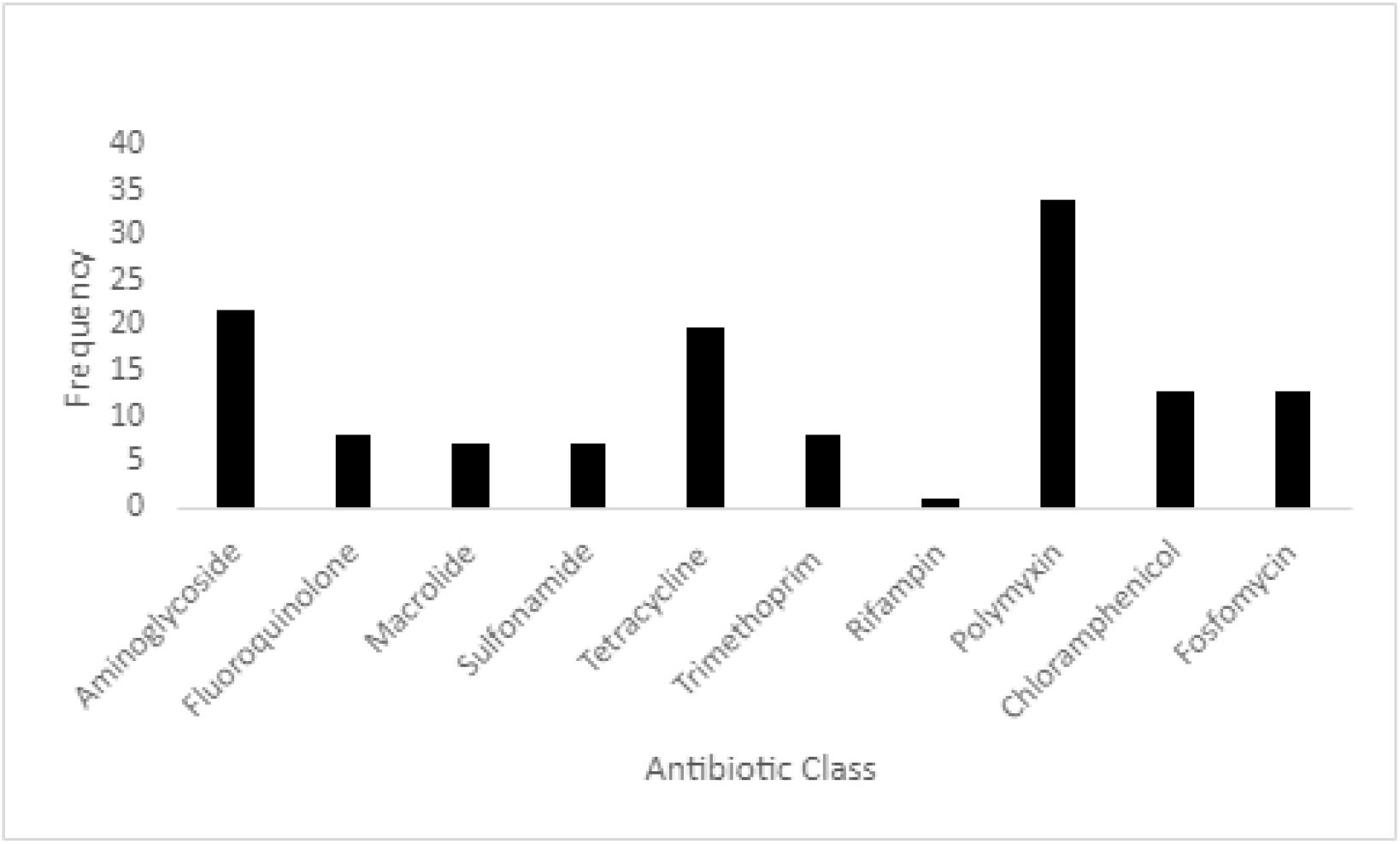
Resistance profile across antibiotic categories. CefR-E isolates were multidrug resistant with most strains carrying genes that confer resistance against polymyxin, aminoglycoside and tetracycline.

## Discussion

This prospective observational study is the first of our knowledge to assess maternal colonization and perinatal transmission of drug-resistant Enterobacterales in the U.S. While studies have reported relatively low rates of colonization among pregnant people with ESBL producing gram-negatives in Western European developed countries (e.g., Germany 2.6%^23^, Norway 2.9%^24^), there are higher rates in the Middle East, Eastern Europe, Asia and Africa including 16.9-21.5% (Israel^25-27^), 19.1% (Lebanon^28^), 12.1-17% (Sri Lanka^29^), 14% (Vietnam^30^), 15% (Africa)^31^; there is a lack of published from the U.S.^32^ Our study found a significant burden of AmpR-E (97%) and CefR-E colonization (14%) in healthy pregnant people and included both urban and suburban patients in Chicago, Illinois.

Since the maternal gut microbiota largely determines the initial microbial population in the neonate, it is imperative to better understand the likelihood of pregnant person colonization with these strains and transmission to infants. Neonatal colonization in our study was 76% with AmpR-E and 7% with CefR-E, with perinatal transmission estimate of CefR-E ranging from 31-50%. Among Gram negative bacteria, Enterobacterales, including *E. coli* and *Klebsiella spp*, are likely causes of neonatal sepsis^33^, and in our study the most common perinatally transmitted CefR-E. The rise in antibiotic resistance among colonizing Enterobacterales, especially against third generation cephalosporins, is very concerning and likely to adversely impact neonatal health at increasing rates even in developed countries such as the U.S.^34-36^. Particularly for preterm infants at high risk of early onset sepsis, approaches to screen for maternal colonization with drug-resistant Enterobacterales to guide empiric antimicrobial therapy or mediate perinatal transmission of such bacteria warrant consideration and further investigation. It is also notable that most pregnant persons were colonized with Enterobacterales resistant to Ampicillin, which is the mainstay of therapy for preterm prolonged rupture of membranes and chorioamnionitis.

Our study attempted to identify clinical factors associated with maternal colonization and perinatal transmission of AmpR-E and CefR-E. Infant nutrition, specifically exclusive breastfeeding versus formula feeding was associated with less likelihood of infant colonization was AmpR-E in the first week of life.

The impact of breastfeeding on infant colonization with maternal drug-resistant bacteria and potential mediation of colonization due to infant nutrition or its attendant alternation of the infant gut microbiome requires further study. Due to relatively limited number of infants with CefR-E and thus limited power, other clinical factors associated with CefR-E colonization may not have been adequately elucidated.

Some interesting observations were noted when CefR-E isolates were sequenced. In the mother-infant dyad NCH0126, we noted that *K. pneumoniae* was isolated from the mother and *E. cloacae* from the infant. However, it is possible that there was co-colonization with both strains in the mother but due to the limitations of detection by bacterial culture only one strain was isolated. In the maternal PWH0104 sample we also noted co-colonization with two different species of Enterobacterales: *P. mirabalis* and *E. coli*. Interestingly, both these species had the same type of beta-lactamases (BlaEC family, CTX-M and Oxa-1). Both CTX-M and OXA ESBLs tend to be plasmid associated which suggests that both these strains may share a plasmid^37,38^. Finally, the mother-infant dyad NCH0104 was colonized with closely related but morphologically variable *E. coli* ST131 strains. On closer analysis both morphotypes had the predicted serotype O25:H4.

Not surprisingly we found that the majority of CefR-E strains isolated from our cohort were *E. coli*, followed by *Enterobacter, Citrobacter* and *Klebsiella*. Interestingly, only three of the 14 *E. coli* isolates did not carry an ESBL gene, but instead had inducible AmpC beta-lactamases. As has been reported elsewhere from regional and international studies we noted that the CTX-M ESBLs, especially CTX-M-15, were the most common ESBLs among *E. coli*^*24,39*^. The presence of these enzymes in isolates from the community cohort is especially worrisome because they are typically associated with conjugative plasmids that are adept in horizontal transfer of genes between Enterobacterales species^40^.

Mother-infant dyads in our cohort carried several *E. coli* sequence types that have been associated with ESBL production as well as early life invasive infections. These include: ST10, ST69 and ST131. In a large international study, which included seven different low- and middle-income countries, these three ST types were found to be the most common *E. coli* subtypes associated with neonatal sepsis^41^. A recent study from the U.S. also showed that ST131 dominated *E. coli* strains associated with neonatal sepsis^42^. Therefore, maternal and infant colonization in this otherwise healthy cohort strongly suggests a community reservoir or these clinically important multi-drug resistant strains.

Among *Enterobacter spp*, ACT/MIR family of beta-lactamases were the most common. These are AmpC-type beta-lactamases that are typically found on chromosomes but more recently have been associated with plasmids^43,44^ as well. Certain plasmid associated sub-types such as *bla*_*MIR-Like*_ *ampC* genes are also found in *Klebsiella spp*, suggesting that they can easily move between different Enterobacterales species^45^. Similarly, the CMH family are part of plasmid mediated AmpC beta-lactamases and are frequently associated with *Enterobacter spp*^*43*^.

All six *Citrobacter spp*. carried the cephalosporinase CMY-4. These CMY-type plasmid-mediated AmpC beta-lactamases are commonly produced by *E. coli* and non-typhoidal *Salmonella*^*46,47*^. Within *Citrobacter spp*, it is thought that CMY-4 may be the precursor of other CMY-type cephalosporinases^48^. Interestingly, at least one other study found CMY-4 within Enterobacterales samples from the community and not from the hospital^46^. The OXY-family of beta-lactamases found in *K. oxytoca* are chromosomally located class A beta-lactamases that at high levels have an ESBL-like cephalosporin hydrolysis profile^49^. These are similar to the OKP-B beta-lactamases in the *K. pneumoniae*^*50*^.

It is important to note that while AmpC beta-lactamases were thought to be largely chromosomal, recent studies point towards a growing prevalence of plasmid associated Amp-C beta-lactamases as well. These have been isolated from across the globe^51,52^, and from varying settings including the community and livestock^53,54^. Given that these enzymes may be both induced and constitutively expressed, exposure to beta-lactam antibiotics may lead to a mixed populations with emergence of high-level resistance against cephalosporins during a treatment course^55^.

The strengths of this study include 1) the prospective, observational investigation of a large cohort of healthy pregnant persons and their infants in a large U.S. city, 2) culture based and WGS providing species and resistance gene data and confidence in perinatal transmission estimates. Limitations include possible drawbacks in rectal swab sampling technique during labor. However, this is limitation would potentially result in underestimation of maternal and infant CefR-E colonization, reinforcing the conclusion that there is significant colonization in healthy pregnant persons in the community, as demonstrated by our sensitivity analyses. Additionally, our study focused on healthy pregnancies with vaginal deliveries. Caesarean deliveries, preterm deliveries, and antibiotic receipt during pregnancy and labor are variables that may impact infant bacterial selection and perinatal transmission, and thus require further study.

In conclusion, pregnant persons in Chicago, a large U.S. city, have significant rates of AmpR-E and CefR-E colonization, similar to high prevalence areas of the world where antimicrobial-resistance is a major concern. Perinatal transmission rates of these drug-resistant bacteria are high, resulting in a significant portion of well infants colonized with resistant Enterobacterales within the first week of life and with plasmid-mediated transmissible resistance genes. Factors such as infant feeding practice warrant future investigation, given the preliminary observation that exclusive breastfeeding is associated with a reduction in perinatal transmission of AmpR-E. If these findings are replicated, surveillance and interventional approaches to mediate early life colonization with drug-resistance Enterobacterales may be indicated.

## Data Availability

All data produced in the present study are available upon reasonable request to the authors.

