## Supplemental Table for "Maternal Colonization, Perinatal Exposure, and Neonatal Acquisition of Resistant Enterobacterales"

| Sample ID | Sample type | Species | Aminoglycoside (gidB and S12p in all) | Fluoroquinolone | Macrolide | Sulfonamide | Tetracycline | Trimethoprim | Rifampin | Polymyxin | Chloramphenicol | Fosfomycin |
|---|---|---|---|---|---|---|---|---|---|---|---|---|
| NCH0002 | M | Enterobacter cloacae | | | | | | | | PmrF | CatA family | FosA2 |
| NCH0009 | M | Enterobacter cloacae | | | | | | | | PmrF | | FosA2 |
| NCH0012 | M | Enterobacter cloacae | APH(6)-IC/APH, APH(3")-I, -Ib, APH(6)-Id, | | | | Tet(B), tetADR | | | PmrF | | FosA2 |
| NCH0016 | M | Escherichia coli | aadA22, aadA, AAC(3)-II,III,IV,VI,VIII, IX,X, APH(3)-IIc, APH(3')-Ia, -I fyuA, | | Mef(B) | | tetC, Tet(A), fyuA | dfrA14 | | | floR, FloR family | |
| NCH0018 | I | Enterobacter cloacae | | | | | | | | PmrF | | FosA2 |
| NCH0023 | M | Escherichia coli | APH(6)-Ic/APH(6)-Id, APH(3")-I, -Ib, APH(3")-Ib, fyuA, | | | sul2 | fyuA, Tet(A), tetC | | | PmrC EF, arnA, mgrB | | |
| NCH0024 | I | Enterobacter cloacae | | | | | | | | PmrF | | Fos2A |
| NCH0031 | M | Escherichia coli | fyuA | | Mph(A) family, Mrx | | fyuA, | | | PmrC EF, arna, mgrB | | |
| NCH0033 | M | Enterobacter cloacae | | | | | | | | PmrF | | Fos2A |
| NCH0047 | M | Escherichia coli | aadA2, AAC(3)-II,III,IV,VI,VIII, IX,X, AAC(3)-IIa, APH(3")-I,-Ib, APH(3')-I,-Ia, APH(6)-Ic/APH(6)-Id, APH(6)-Id, | QnrB family, QnrS1 | Mph(A) family, Mrx | sul1, sul2 | Tet(A)(B), tetCR, | dfrA12 | | PmrC F, arnA, mgrB | CatA1, catI | |
| NCH0049 | M | Escherichia coli | fyuA, | | | | fyuA | | | PmrC EF, arnA, mgrB | | |
| NCH0076 | M | Escherichia coli | aadA22, AAC(3)-II,III,IV,VI,VIII, IX,X,AAC(3)-IIa , APH(3")-I,-Ib, APH(3')-I, -Ia, APH(6)-Ic/APH(6)-Id, APH(6)-Id | QnrB family, QnrS1, | Mef(B), Mph(A) family, Mrx | sul3 | Tet(A) | dfrA14 | arr-2 | PmrC F, arnA, mgrB | ClmA family, FloR family, floR | |
| NCH0104 | I (pink colonies) | Escherichia coli | fyuA, AAC(6')-Ib-cr, | | | | fyuA | | | PmrC EF, arnA, mgrB | CatB family, catB3 | |

| | | | | | | | | | | | | |
|---|---|---|---|---|---|---|---|---|---|---|---|---|
| NCH0104 | M (pink colonies) | Escherichia coli | fyuA, AAC(6') -lb-cr, | | | | fyuA | | | PmrC EF, arnA, mgrB | CatB family, catB3 | |
| NCH0104 | I (white colonies) | Escherichia coli | fyuA, AAC(6') -lb-cr, | | | | fyuA | | | PmrC EF, arnA, mgrB | CatB family, catB3 | |
| NCH0104 | R (while colonies) | Escherichia coli | fyuA, AAC(6') -lb-cr, | | | | fyuA | | | PmrC E, arnA, mgrB | CatB family, catB3 | |
| NCH0112 | M | Escherichia coli | fyuA, | QnrB10, QnrB4 | Mph(A ) family, Mrx | sul1 | fyuA | dfrA7 | | PmrC EF, arnA, mgrB | | |
| NCH0112 | I | Escherichia coli | fyuA, | QnrB10, QnrB4 | Mph(A ) family, Mrx | sul1 | fyuA | dfrA7 | | PmrC EF, arnA, mgrB | | |
| NCH0126 | I | Enterobacter cloacae | fyuA | | | | fyuA | | | PmrF | | FosA2 |
| NCH0126 | M | Klebsiella pneumoniae | | | | | | | | PmrF | | FosA5 |
| NCH0136 | I | Citrobacter Freundii | | QnrB10, QnrB6 | | | | | | PmrB CEF, arnA | | |
| NCH0137 | I | Citrobacter spp | | | | | | | | PmrB CF, arnA | | |
| NCH0138 | I | Escherichia coli | aadA5, AAC(3)-II,III,IV,VI,VIII, IX,X, AAC(3)-IIa, AAC(6')-Ib-cr, APH(6)-Ic/APH(6)-Id, APH(3'')-I,-Ib, APH(6)-Id | QnrB family, QnrS1 | | sul1, sul2 | tetAR, Tet(B) | dfrA17 | | PmrC F, mgrB, arnA | CatB family, catB3 | |
| NCH0141 | M | Escherichia coli | AAC(3)-II,III,IV,VI,VIII, IX,X, APH(3')-I,-Ia, AAC(3)-IIc, fyuA | | | | fyuA | | | PmrC EF, arnA, mgrB | | |
| PWH0017 | I | Enterobacter cloacae | | | | | | | | PmrF | | FosA2 |
| PWH0025 | M | Escherichia coli | AAC(3)-II,III,IV,VI,VIII, IX,X, AAC(3)-IIc, AAC(6')-Ib-cr, fyuA, | | | | fyuA | | | PmrC EF, arnA, mgrB | CatB family catB3 | |
| PWH0028 | M | Escherichia coli | | | | | | | | PmrC F, arnA, mgrB | | |
| PWH0031 | I | Escherichia coli | | | | | | | | PmrC F, arnA, mgrB | | |
| PWH0031 | M | Escherichia coli | fyuA | | | | fyuA | | | PmrF, arnA, mgrB | | |
| PWH0041 | | Enterobacter cloacae | | | | | | | | | CatA1/CatA4 family | FosA2 |

| | | | | | | | | | | | | |
|---|---|---|---|---|---|---|---|---|---|---|---|---|
| PWH0050 | I | Klebsiella Oxytoca | | | | | | | | | | FosA5 |
| PWH0060 | M | Citrobacter Freundii | AAC(6')-lc,f,g,h,j,k, l, r-zAAC(6')-lf | QnrB10 | | | | | | PmrB CF, arnA | | |
| PWH0062 | M | Citrobacter spp | | | | | | | | PmrB CF, arnA | | |
| PWH0073 | I | Klebsiella aerogenes | | | | | | | | PmrF | | FosA5 |
| PWH0074 | M | Citrobacter spp | | | | | | | | PmrB CF. mgrB | | |
| PWH0086 | M | Citrobacter Freundii | APH(3")-l,lb, APH(6)-lc/ld | QnrB10 | | | | | | PmrB CF, arnA, mgrB | | |
| PWH0094 | M | Enterobacter cloacae | | | | | | | | | | FosA2 |
| PWH0104 | M (white colonies) | Proteus mirabalis | AAC(6')-lb-cr, aadA | | | | Tet(J) | dfrA1 | | | CatA1/CatA4, CatB family, catB3 | |
| PWH0104 | M (pink colonies) | Escherichia coli | AAC(6')-lb-cr, aadA | | Mph(A) family, MrX | sul1,2 | Tet(b), tetADR | dfrA1,17 | | PmrB CEF, arnA | CatB family | |

Genotypic resistance profile of CefR-E strains isolated in this study for non-beta-lactam antibiotic classes. M - maternal, I – infant. Blue indicates the non-beta lactam antibodies and the resistance genes found.
